# A study protocol on assessing the acceptance and effectiveness of a digital adherence technology for TB preventive treatment in Bangladesh

**DOI:** 10.1101/2025.04.23.25326267

**Authors:** Pushpita Samina, Md Rifat Haidar, Tasmia Ibrahim, Tanjina Rahman, Mohammad Shahnewaz Sarker, Syada Samsia Noor, Shahriar Ahmed, Mohammad Khaja Mafij Uddin, Senjuti Kabir, Sayera Banu

## Abstract

**Background:** Adherence to tuberculosis preventive treatment (TPT) remains a significant challenge in high-burden countries like Bangladesh, where approximately 44 million people are infected with latent tuberculosis.

**Methods/Design:** This study protocol describes a mixed-methods observational study to evaluate “iDOTS,” a locally developed digital adherence technology adapted from 99DOTS, for monitoring and improving TPT adherence among adult household contacts of bacteriologically confirmed pulmonary TB patients. The study will be conducted in two districts in Bangladesh with similar geographical and societal characteristics, with Narsingdi as the intervention site and Manikganj as the control site. Applying the Unified Theory of Acceptance and Use of Technology (UTAUT), we will assess technology acceptance, implementation challenges, and effectiveness through quantitative and qualitative approaches. The quantitative component will compare TPT adherence between iDOTS users and non-users, while qualitative interviews will explore user experiences and attitudes among healthcare providers and patients. Adherence will be verified through a combination of digital records, self-reports, and random isoniazid urine testing.

**Discussion:** With an estimated sample size of 422 patients and 77 healthcare providers, this study aims to generate evidence that if digital adherence technologies can strengthen TPT implementation in resource-limited settings. The findings will address critical gaps in the TPT cascade and inform strategies for scaling up TPT nationally, ultimately supporting global efforts to reduce the TB disease burden through effective preventive measures.

**Trial registration:** ‘Not applicable’

## Background

The World Health Organization (WHO) defines latent tuberculosis infection (LTBI) as a state of persistent immune response to *Mycobacterium tuberculosis* antigens without any clinically manifested active tuberculosis (TB) [1]. Approximately one-quarter of the global population is estimated to be infected with latent TB infection. This latent phase can persist for a lifetime in otherwise healthy individuals. Reactivation occurs in about 5% to 15% of individuals, typically within two to five years following the initial infection. This reactivation process transforms a latent infection into active TB disease, thereby making individuals with latent TB infection a significant reservoir for new active tuberculosis cases [1]. To reduce the burden of TB disease, the WHO recommends the use of TB preventive treatment for the high-risk and close contact of bacteriologically positive pulmonary TB patients with latent TB infection. The recent TB preventive treatment guideline from the WHO recommends three shorter treatment regimens for latent TB infection treatment [2]. Despite being termed “shorter”, these regimens require a minimum of three months of treatment, including three once-a-week doses of isoniazid and rifapentine (3HP), three daily doses of isoniazid and rifampicin (3HR), and four months of daily rifampicin (4R). The success in preventing TB disease and averting TB infection to diseases significantly depends on treatment adherence and completion. Moreover, as individuals with latent TB infection do not exhibit any clinical manifestations of TB disease, motivating healthy individuals to complete a 3-month or 4-month treatment regimen presents challenges at the implementation level [3, 4].

In addition, all member states are mandated by the WHO to report on the TB preventive treatment cascade, which includes data on the number of eligible populations, the number of populations under evaluation, the number of contacts with latent TB infection, the number of individuals starting treatment, and the number of individuals completing treatment. Since the implementation of TB preventive treatment for non-HIV-infected adult individuals is relatively new, the reporting systems are still not well-equipped in many countries with a high burden of TB. The reporting systems often fail to capture the adverse drug reactions among individuals undergoing TB preventive treatment. Therefore, there is a need to develop a reporting system that can encourage individuals to adhere to TB preventive treatment and assist in reporting adverse events and the treatment outcome.

Bangladesh is one of the countries with a high burden of TB, and it is estimated that approximately 26.7% of the total population (∼44 million) is infected with latent TB infection [5]. Bangladesh started implementing TB preventive treatment for adult household contacts in 30 districts (out of 64 districts) in 2022 and gradually expanded nationwide. However, the country lacks a robust reporting system for overseeing TB preventive treatment adherence and completion. The National TB Control Program (NTP) solely focuses on self-reported treatment completion, and there is an absence of a structured mechanism to support individuals in medication adherence, monitor their compliance, or report any adverse events [6].

Various technological interventions, including digital adherence technology, have been developed to monitor adherence to TB treatment [7]. One of the utilized technologies is “99DOTS”, a low-cost, cellphone-based digital adherence technology for TB treatment [8, 9]. While multiple studies have appraised the use of “99DOTS”, most of these have concentrated on adherence to standard TB treatment [9, 10]. Research examining the adherence monitoring of TB preventive treatment through “99DOTS” is limited, primarily focusing on specific high-risk populations such as HIV/AIDS patients and pediatric contacts [11-13]. Limited evidence exists regarding the utilization of “99DOTS” or other digital adherence technology for TB preventive treatment in non-HIV/AIDS adult contacts.

Drawing from the foundation of 99DOTS, the International Centre for Diarrhoeal Disease Research, Bangladesh (icddr,b), has developed a digital adherence technology for TB preventive treatment named “iDOTS.” Its principal aim is to facilitate real-time monitoring of TB preventive treatment-enrolled contacts. Through the “iDOTS” dashboard, community health workers or medical assistants can supervise individual contacts as they take their medication. Each contact is required to register with “iDOTS” and receive a colored pouch containing medication and a printed phone number. Subsequently, the contacts are mandated to call a specific toll-free number to confirm medication intake, and upon completion, they receive a confirmation SMS from the server. This technology empowers community health workers to identify non-adherent patients in real-time and provide follow-up care promptly.

The proposed protocol seeks to evaluate the utilization of “iDOTS” to determine its acceptance and effectiveness in enhancing TB preventive treatment outcomes in Bangladesh. The objective is to improve the adherence and outcome of TB preventive treatment among the eligible close contacts of bacteriologically confirmed pulmonary TB patients through remote monitoring of real-time adherence records. The protocol outlines how quantitative and qualitative data will be utilized to inform the acceptance, implementation, and effectiveness of “iDOTS” in improving TB preventive treatment adherence in Bangladesh.

## Methods/Design

### Study objectives

The primary objective of this study is to evaluate the role of “iDOTS” in the implementation of TB preventive treatment in Bangladesh. The specific objectives are as follows:

1. To explore how the attitudes of users (both providers and beneficiaries) influence the acceptance of “iDOTS” for adherence to TB preventive treatment in Bangladesh.
2. To assess the effectiveness of “iDOTS” in maintaining adherence and reporting on TB preventive treatment at the individual level.
3. To document users’ experiences and the benefits and challenges encountered while implementing “iDOTS.”

### Study design

We have applied the “User Acceptance of Information Technology: Toward a Unified View Theory” (UTAUT) to design this study. It is a modern theory that provides a helpful tool for managers to understand the drivers of acceptance of a new information technology used in health. It also helps assess the likelihood of success for new technology introduction and design interventions (including training, marketing, etc.) targeted toward users who are less interested in adopting new technology. The theory identifies four variables influencing the “behavioral intention” of using a new technology [14]: performance expectancy, effort expectancy, social influence, and facilitating conditions. In addition, during a pragmatic trial of e-health interventions, evidence should also be collected on the organization, cost, social, human, technological, clinical, legal, and ethical aspects [15].

We propose that our study be a mixed-method observation study, where we plan to use quantitative data to collect evidence on the technological and clinical aspects and qualitative data to get detailed information on the social and human aspects. With this study, we want to compare the adherence to TB preventive treatment between the two groups, users and non-users of “iDOTS” and capture the user experience before, during, and after the intervention. We aim to collect both quantitative and qualitative data through pre- and post-intervention exit interviews, which will help predict the acceptance of “iDOTS”. The quantitative analysis will help to understand the variation in the pattern of acceptance with age, sex, and education. However, the quantitative analysis alone cannot explain the causes behind the variation unless detailed information is collected through user interviews. The qualitative study will help understand the challenges and identify possible solutions to the quantitative study’s findings. In addition, we want to capture users’ (both healthcare providers and individuals receiving TB preventive treatment) experience that will supplement the effectiveness results. In a scenario where the effectiveness is high, but the users are not satisfied, it will be challenging to scale up the technology unless their issues are addressed. We also want to assess if the technology alone is a compelling reminder or if they use any other method to remind themselves about the medicine. An in-depth interview with the users will also give information on the quality of “iDOTS” and the confidence and comfort of providers and patients in using “iDOTS”. We will collect qualitative and quantitative data concurrently to assess if the results of both data converge or diverge.

### Study setting

The adult household contacts are enrolled for TB preventive treatment through Upazila health complexes and district hospitals. Considering the implementation challenge and ethical constraints, we predicted that randomly assigning individuals to the intervention group would be challenging. While cluster randomization could have been a possible solution, the NTP in Bangladesh had a preference for an intervention district; based on their preference, we purposively selected the Narsigndi district as an intervention site. Considering the geographical and societal similarities, we selected the Manikganj district as a control site. Six directly observed treatment (DOT) centers placed at Upazila health complexes of Raipura, Monohardi, Belabo, Palash, Shibpur, and Sadar, and four DOT centres placed at Upazila health complexes of Ghior, Singair, Shibaloy, and Sadar hospital in Manikganj has been included as study sites.

### Sample size

Sample size for quantitative data collection

#### The sample size for end-users

Considering the district demography, latent TB infection burden, and generalizability, we calculated the sample size as below:

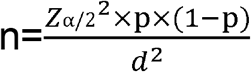

Where,

Zα/2 = 1.96 (Considering 95% confidence level and α is 0.05)

p= 0.50 (Proportion)

d= 0.05 (Precision of the estimate)

Considering data from our previous study on monitoring adherence to the standard TB treatment using a digital platform (unpublished data), we assume that approximately 50% of the TB preventive treatment-initiated individuals will consent to enroll for digital adherence. Based on the above formula, considering 5% absolute precision, a total of 384 participants will be needed. Considering a 10% refusal rate, the required sample size will be 422 to determine the adherence and outcome of TB preventive treatment among the eligible household contacts of bacteriologically confirmed pulmonary TB patients. We plan to sequentially enroll and survey all eligible participants identified during the screening activity from intervention sites. Recruitment will continue until we have identified 422 from intervention sites.

#### The sample size for healthcare providers

For the survey, the following Cochrane’s formula is used to determine the required sample size

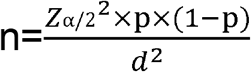

Where,

Zα/2 = 1.96 (Considering 95% confidence level and α is 0.05)

p= 0.95 (Proportion)

d= 0.05 (Precision of the estimate)

The sample size assumes that 95% of healthcare providers reported that they had previous experience using a smartphone in a digital adherence technology evaluation study [16]. Based on Cochrane’s formula, considering 5% absolute precision, a total of 73 healthcare providers will be needed, and considering a 5% refusal rate, the required sample size will be 77 healthcare providers to estimate the experiences issues of using “iDOTS”.

#### Sample size for qualitative data collection

Qualitative interviews will be conducted with both end users and service providers prior to and following the intervention. Data collection will continue until saturation is achieved, ensuring a comprehensive understanding of participant experiences. Simultaneous data coding will be employed to monitor and identify the point at which saturation occurs, allowing for a rigorous and systematic analysis of the interview data.

### Participant enrollment

The baseline data on TB preventive treatment eligibility in the Narsingdi district exhibited that in 2023, around 2800 individuals were enrolled for TB preventive treatment over 12 12-month period. Considering our 99DOTS pilot study, we assume that approximately 50% of the TB preventive treatment-initiated individuals will consent to enroll for digital adherence, which would be 1400. During the six-month enrolment period and considering enrolment criteria (90% eligibility), we could enroll 1400/2 = 700 X 85% = 595, i.e., approximately 600 eligible TB preventive treatment individuals. We will also enroll 600 participants as a control from another site (Manikganj) following the same criteria.

### Participant selection

For the intervention group end users, all eligible adult (> 18 years) household contacts of people with bacteriologically positive pulmonary TB will be enrolled in the study following the national TB preventive treatment algorithm (Fig 1). The community healthcare workers will screen the eligible household contacts of people with TB (bacteriologically positive) for TB symptoms (chronic cough, fever, weight loss). The individuals with symptoms will be referred to facilities, and the physicians will evaluate them for TB disease with chest X-ray and GeneXpert. The asymptomatic individuals will be given a tuberculin skin test, and if the test results are positive, they will be further evaluated with an X-ray. If there are changes in the X-ray, the individuals will be evaluated for TB disease and enroll for the TB treatment. The ones who do not have changes in chest X-ray are enrolled for TB preventive treatment. The end-user participants for the control group will be enrolled following the same process.

**Fig 1.**
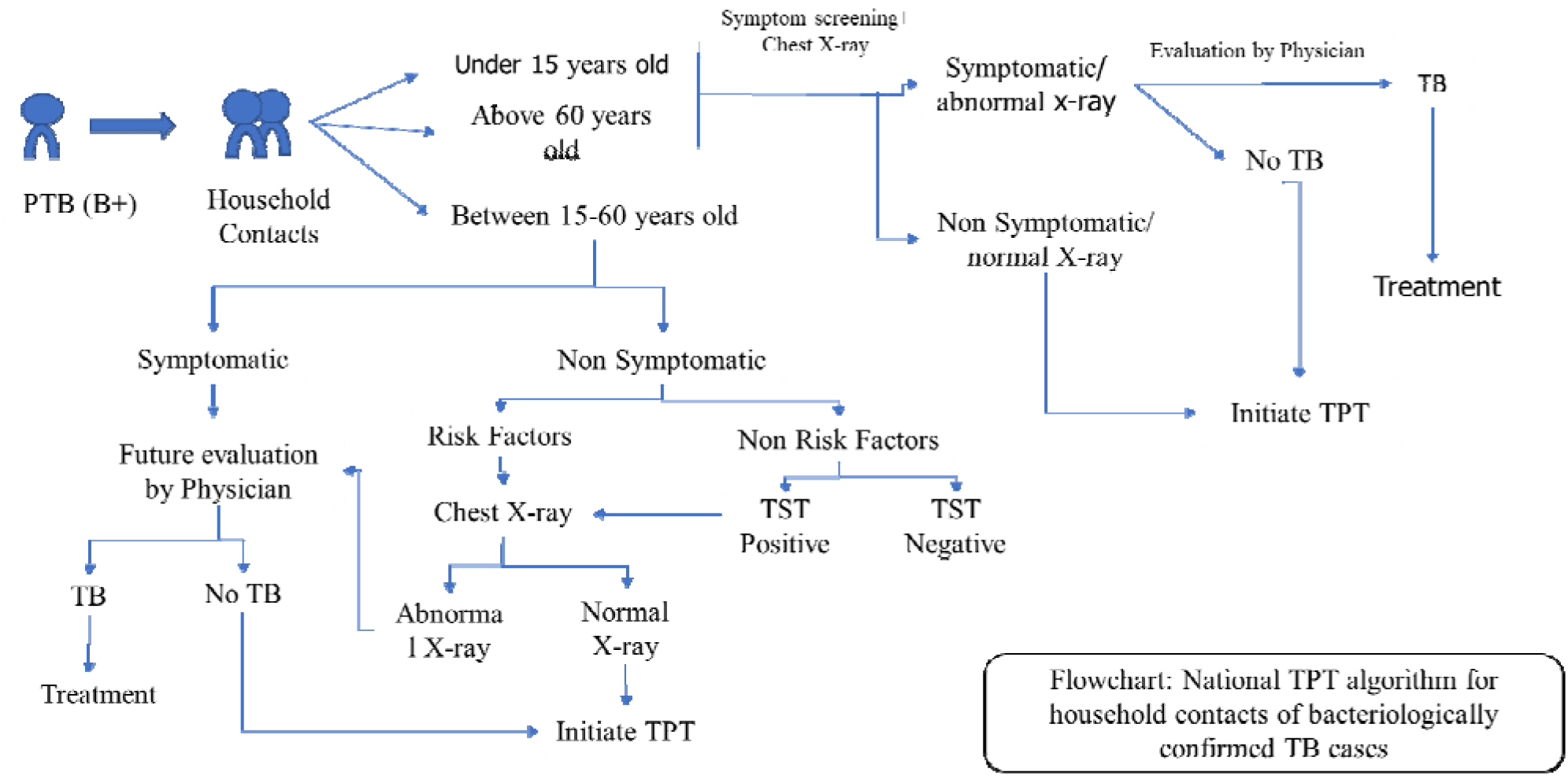
TPT algorithm based on national TB control programme

For another group of users, healthcare providers, we will select the providers who are directly involved in the implementation process of TB preventive treatment, including community healthcare workers, medical technologists, TB and leprosy control assistants, physicians, and healthcare managers (upazila health and family planning officer) from the intervention group.

### Implementation process

The study will have three phases. (Fig 2) illustrates the step-by-step process. In the first phase (pre-intervention), the study assesses the users’ attitudes toward “iDOTS”. The second phase is the implementation phase, and in the third phase (post-intervention), the study will capture the outcomes and user experiences. Data are collected from the users using demographic information questionnaires after obtaining informed written consent in the first phase and stored in an in-house secured locker. We conduct in-depth interviews with a selected group of individuals with semi-structured questions. The questionnaires are focused on assessing users’ attitudes towards “iDOTS”. They are briefed about “iDOTS” prior to the interviews. All interviews are conducted in Bangla and last for 30-45 minutes. The interviews are recorded, transcribed, and translated into English. Users of “iDOTS” also receive a document with instructions and a manual form to track the doses.

**Fig 2.**
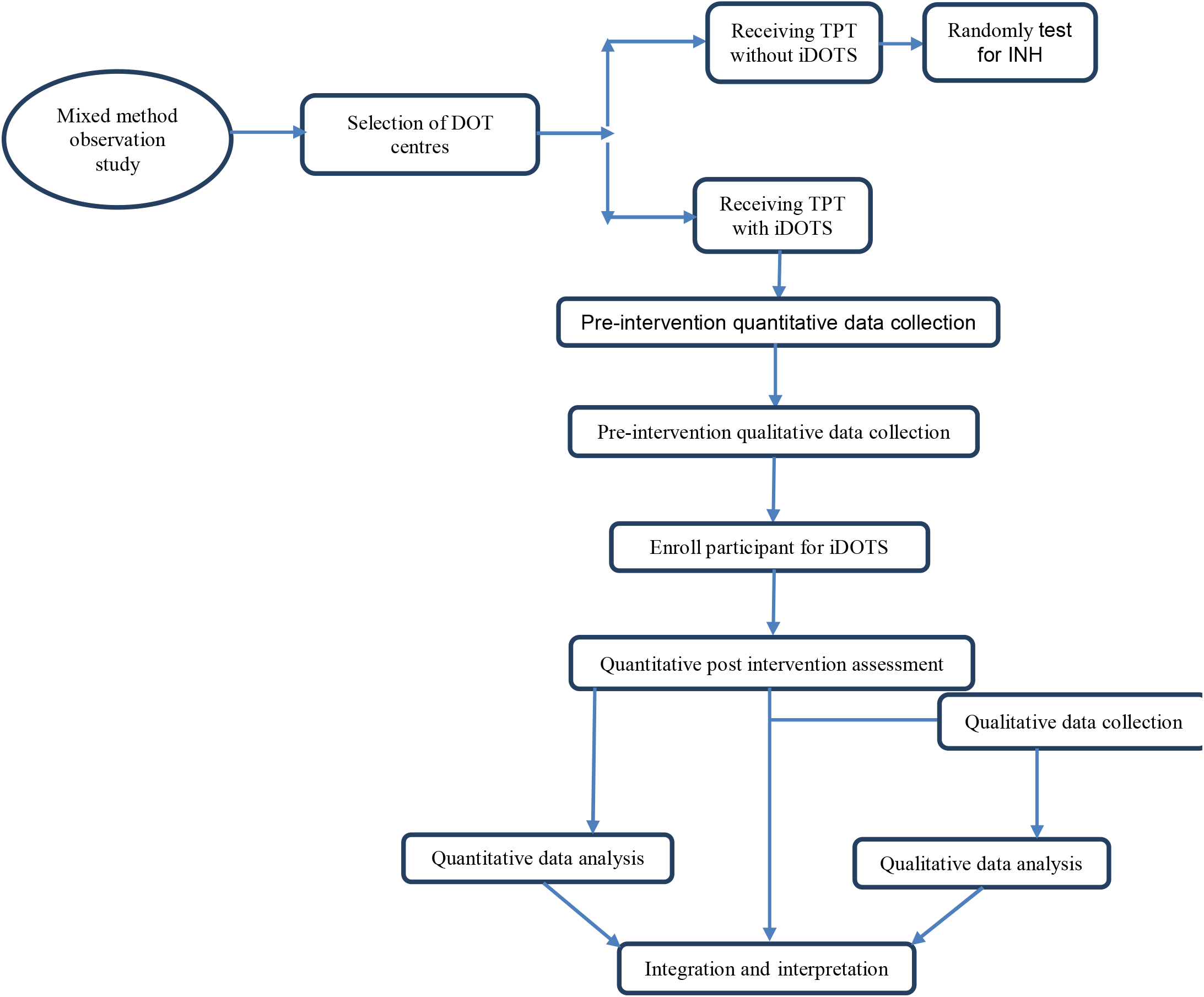
diagram of iDOTS study design

In the intervention group, the end users are enrolled for “iDOTS” and are given a document with instructions and a manual form to track the dose. The “iDOTS” contains TB preventive treatment medication in tailored colorful pouches containing a printed phone number on it. The contacts are instructed to call the specific toll-free numbers from their pre-registered number each time after taking the medication. The dialed call reaches a predesigned automated call receiving system. It is linked to a well-designed tracking algorithm that marks each enrolled participant’s status as medication taken or skipped.

The medical technologist (MTs) who enroll and counsel the TB patients will check the “iDOTS” dashboard designed for him/her every morning. If any individual misses a dose, the technical assistant calls him /her to remind about the missing dose and counsels to complete the treatment. In addition, a medication reminder SMS will be sent to the participants from whom no call was received on a particular day from the specified number until 10.00 PM at night.

To assess the accuracy of the call and adherence, we randomly test the participant’s urine using a strip-based testing (IsoScreen assay) kit to examine the presence of isoniazid (a TB preventive treatment drug). We maintain a calendar to randomly select the individuals for urine tests and ensure that each participant is tested at least once during their full course of treatment. The informed consent taken during the enrolment includes the information on the urine test. The urine is collected with a sterile container at the participants’ home and tested with the strip on the spot. The test results are recorded digitally.

For the control group, a similar measure will be taken to measure and observe the adherence. Individuals who enrolled for TB preventive treatment will be randomly assigned for urine tests, and urine will be collected at their home and tested and recorded on the spot.

The medical technologists and community healthcare workers were trained centrally on project activities, digital adherence monitoring on the “iDOTS” platform, patient enrollment, and counseling. The project research officer alternately visits the facilities and supervises the activities of medical technologists and community healthcare workers. One to two DOTS providers from each DOTS facility were sensitized at the district level on the “iDOTS” platform, patient enrollment, and counseling for understanding and supporting the overall activity.

In the study’s second phase, we will use the data from the “iDOTS” database to measure the outcomes (adherence to TB preventive treatment and missed doses). Concurrently, we will interview the same individuals to collect qualitative data from the providers and patients using a semi-structured questionnaire. This part of the study wants to capture the human experiences to augment the findings from the experiment. The providers and patients will be interviewed to discuss their experiences and to capture the benefits and challenges of “iDOTS.”

### Data management

This is a complex study with multiple components. All data will be collected with an electronic tablet and saved in a cloud-based system. Research assistants will conduct the interviews for the qualitative data collection. All interviews will be recorded with the participant’s consent. We will ensure privacy throughout the data collection and storage, as per the requirements of the ethical review board.

### Data analysis

The data analysis will be performed with the aim of assessing the effectiveness of “iDOTS”, to identify the variables that influence the uptake of this new technology, “iDOTS” along with documentation of users’ experience.

### Quantitative data analysis

To assess the effectiveness of “iDOTS”, we will calculate the adherence rate to TB preventive treatment, measured through multiple data sources: “iDOTS” dashboard data, self-reported adherence, manually collected information, and urine sample iso-screen test results. Adherence will be calculated as the percentage of doses taken relative to the total prescribed, based on the “iDOTS” dashboard data. In addition, self-reported adherence will be cross-checked with the urine sample results to assess self-reports’ accuracy. The data from the intervention group (using “iDOTS”) will be compared to the control group (using traditional methods) to determine the effectiveness of the “iDOTS” system in improving adherence.

To assess differences in adherence between groups, we will first compute descriptive statistics, including the mean, median, and standard deviation, to summarize the adherence rates in both groups. Differences between the control and intervention groups will be evaluated using the appropriate statistical tests, such as the Mann-Whitney U test for non-parametric data or the independent t-test for normally distributed data. The focus will be on the median number of missed doses, as well as the variance between self-reported adherence and adherence confirmed by urine sample results.

In addition to basic group comparisons, regression analyses will be employed to identify factors influencing adherence. Multiple linear regression will be used for continuous outcomes (e.g., adherence rates), while logistic regression will be applied to examine binary outcomes, such as whether a participant is adherent or non-adherent. These regression models will help identify demographic and clinical variables that predict adherence and allow for the identification of key factors that may influence the effectiveness of the “iDOTS” technology.

Further, Partial Least Squares (PLS) regression will be applied to explore the relationships between multiple predictors, including demographic factors, attitudes toward the technology, and UTAUT constructs (performance expectancy, effort expectancy, social influence, and facilitating conditions). PLS path modelling will allow us to assess these constructs’ direct and indirect effects on adherence behaviour. For instance, the analyses will investigate how users’ perceptions of the ease of use and perceived effectiveness of “iDOTS” (effort expectancy and performance expectancy) influence their adherence to TB preventive treatment. Similarly, the role of social influence (e.g., support from healthcare providers, family, or peers) and facilitating conditions (e.g., availability of smartphones and technical support) will be evaluated for their impact on technology uptake and adherence.

### Qualitative Analysis

The qualitative component will focus on capturing users’ experiences, attitudes, and perceptions of using “iDOTS”. This will include exploring their satisfaction with the technology, challenges faced during its use, and the broader social and logistical factors that affect adoption. Semi-structured interviews will be conducted with participants to understand the nuanced factors influencing their engagement with “iDOTS”.

The qualitative data will be analyzed using thematic analysis, which involves identifying and interpreting patterns (or themes) within the data. This process will include familiarization with the data, generating initial codes, searching for themes, reviewing and refining themes, defining and naming themes, and reporting the findings. Key themes will likely emerge around areas such as ease of use, trust in the technology, perceived benefits and barriers, and external factors such as technical support and social influence. These qualitative themes will provide deeper insight into the quantitative findings, particularly in terms of why certain demographic groups exhibit lower adherence rates or experience challenges with the technology.

### Integrating Quantitative and Qualitative Data

The integration of quantitative and qualitative data will provide a comprehensive understanding of the effectiveness of “iDOTS” and the factors that influence its uptake. The findings from both datasets will be merged through a process known as **data triangulation**, allowing for a more nuanced interpretation of the results. The combined analysis will enable us to assess how user experiences (captured qualitatively) explain or complement the patterns observed in the quantitative data.

For instance, if the quantitative data indicates that adherence rates are significantly higher in specific groups, qualitative data will be used to explore the reasons behind this success, such as user satisfaction, ease of use, or strong social support. Conversely, if certain groups show lower adherence, qualitative insights will help pinpoint the challenges they face—such as technological difficulties, lack of awareness, or social stigma—and suggest ways to overcome these barriers. This integration of findings will create a richer and more holistic picture of “iDOTS” impact on TPT adherence.

The joint display of quantitative and qualitative results will facilitate the comparison of adherence patterns with the underlying user experiences. For example, comparing adherence rates in different demographic groups can be placed alongside thematic insights related to trust, ease of use, or barriers to adoption. This will allow for a more direct comparison between how users experience the technology and its impact on their adherence behavior. It will provide insights into whether the technology is a scalable and effective solution for improving TB preventive treatment adherence in Bangladesh.

## Discussion

The implementation of TB preventive treatment remains a relatively new and challenging territory for many low- and middle-income countries, particularly those burdened with high rates of TB, like Bangladesh. While TB preventive treatment implementation has gained momentum in countries with high HIV/AIDS prevalence, where it is easier to integrate into existing healthcare systems through platforms that track and counsel individuals, the situation in Bangladesh presents unique challenges. In countries like Bangladesh, where healthcare infrastructure is still developing, scaling TB preventive treatment across a large population becomes more complicated, requiring technological solutions and systemic changes in healthcare delivery.

It has been over five years since the United Nations High-Level Meeting on TB, where member states committed to implementing TB preventive treatment nationally [17]. However, data from various countries, including Bangladesh, indicate that progress has been slow. While the COVID-19 pandemic is often cited as a major disruptor to TB services, and the high cost of shorter drug regimens is a significant financial barrier, there are also other underlying issues hindering the large-scale implementation of TB preventive treatment. A key factor contributing to this slow pace of implementation is the lack of sufficient tools and evidence that can guide and support countries in scaling up their TB preventive treatment efforts [18].

In Bangladesh, where the need for innovative solutions is critical, locally developed tools such as the “iDOTS” system can potentially fill some of the gaps in the existing healthcare infrastructure. However, while the effectiveness of such tools is still largely unknown, this tool, “iDOTS” could serve as a valuable resource for improving the monitoring and reporting of TB preventive treatment adherence. By enabling the remote monitoring of medication intake, “iDOTS” could provide real-time data on treatment adherence, identify gaps in care, and help reduce the risk of treatment failure. In this context, “iDOTS” could serve as an adherence tool and a mechanism for identifying adverse events and monitoring treatment progress.

Despite the potential benefits, adopting such a locally developed technological solution is not without barriers for most low-middle income countries. Bangladesh’s diverse healthcare landscape, coupled with varying levels of technological infrastructure and literacy, presents a set of challenges to the widespread uptake of digital tools like “iDOTS”. Although the technology is relatively simple, its acceptance and integration into the daily lives of healthcare providers and patients may be hindered by issues such as limited access to smartphones, internet connectivity problems, and resistance to adopting new technologies in rural areas. Additionally, healthcare workers lack awareness and training, which could limit their ability to leverage such tools effectively.

The UTAUT (Unified Theory of Acceptance and Use of Technology) provides a useful framework for understanding technology acceptance. Still, it accounts for only about seventy percent of the variance in user acceptance [14]. This suggests that other factors at play influence the decision to adopt and use technologies like “iDOTS”. The first phase of this mixed-methods study aims to identify additional constructs beyond those specified by UTAUT, which could help improve predictions of technology adoption and usage behaviour. Qualitative interviews can help us identify such variables. By incorporating these additional factors into the UTAUT framework, this study will contribute empirical evidence to refine and expand the theory, particularly in TB treatment in low-resource settings.

One of the most pressing issues in the implementation of TB preventive treatment is the uncertainty around whether people are taking their medication consistently and whether counselling efforts are adequate. Despite the substantial resources allocated to TB preventive treatment, there is still a significant knowledge gap regarding the actual number of people taking the medicine, the rate of treatment dropouts, and whether the preventive therapy is achieving the intended public health impact. “iDOTS” could potentially bridge this gap by providing more accurate and timely data on treatment adherence and by offering a platform for continuous counselling and support. However, to fully understand its potential, further studies are needed to assess its effectiveness in real-world settings, particularly in countries with limited healthcare infrastructure like Bangladesh.

Moreover, as we begin to integrate such technologies into TB programs, it is critical to monitor the adherence rates and the broader social and contextual factors that influence treatment uptake. This includes the level of trust patients have in the healthcare system, the perceived stigma associated with TB, and the adequacy of healthcare resources available for remote consultations. It is essential that any technological intervention also be accompanied by efforts to address these broader barriers to care in order to achieve meaningful improvements in public health outcomes.

While “iDOTS” represents a promising tool for enhancing TB preventive treatment implementation in Bangladesh, its effectiveness in improving adherence will ultimately depend on how well it can be integrated into the existing healthcare system. The evidence generated from this study could serve as a foundation for scaling up this tool, providing a critical resource for monitoring TB treatment adherence, and ultimately improving patient outcomes. However, as we move forward, it is important to recognize that the journey towards large-scale TB preventive treatment implementation is complex and requires multifaceted solutions, including but not limited to technological innovations like “iDOTS”.

Ultimately, this study aims to assess the potential of “iDOTS” in improving adherence and contribute to the broader conversation about how low- and middle-income countries can better implement TB preventive treatment. The findings from this study will provide valuable insights into the barriers and enablers of technology adoption in healthcare settings, particularly in contexts like Bangladesh, where both technological infrastructure and public health systems are still evolving. By refining the UTAUT framework and incorporating additional contextual factors, this research will help pave the way for more effective and scalable TB prevention efforts in high-burden countries.

### Timeline

This study, excluding the product and protocol development phases, will span nine months, featuring a six-month enrollment period followed by a three-month follow-up, data analysis, and manuscript preparation stage. Based on our enrollment rate, we anticipate completing enrollment by May-June 2025. Following this, there will be a three-month follow-up period to monitor adherence to TB preventative treatment medication intake, accompanied by data analysis and manuscript preparation. While we have successfully enrolled notable participants at the intervention site and collected pre-intervention quantitative and qualitative data from both providers and participants, post-intervention data collection will commence at the end of May 2025 and is expected to be finalized by the end of June 2025. Data management and analysis will occur concurrently, with anticipated results being available by July 2025.

## Supporting information

Supporting docs_Manuscript of iDOTS protocol

## Data Availability

The data will be collected digitally through data collecion tab and recorder will be used with proper consent and will be stored in a cloud based sever. The data mangement team and the statistician will manage data. The principal investigator and the study team wil have access to the data. After project's completion, data will not be availble publicly. If data is reuired to be shared, this wil be done as per icddr,b's data sharing policy. Only the deidentifed data will be shared

## List of abbreviations

NTP: National Tuberculosis Programme

DOT: Directly observed treatment

TB: Tuberculosis

TPT: TB preventative treatment

UTAUT: Unified Theory of Acceptance and Use of Technology.

## Declarations

### Ethics approval and consent to participate

The protocol (PR-23001, version: 1.7, January 29, 2025) has been approved by icddr,b’s Institutional Review Board, which includes both the Research Review Committee and the Ethical Review Committee. We have obtained permission from local health authorities and facilities, supported by a letter from the NTP for this project. Project staffs explain the study’s purpose to participants and obtain their informed written consent. We will strictly maintain the confidentiality of participants regarding their health status and ensure data anonymity. Access to anonymous data will be limited to authorized health personnel, implementers, and participants.

### Consent for publication

‘Not applicable’

### Availability of data and materials

The data will be collected digitally through data collection tab and store in a cloud based sever. The data management team and the statistician will manage data. The principal investigator and the study team will have access to the data. After project’s completion, data will not be available publicly. If data is required to be shared, this will be done as per icddr,b’s data sharing policy. Deidentified research data will be made publicly available when the study is completed and published.

## Authors’ contributions

Pushpita Samina

Roles: conceptualization, supervision, writing – original draft, editing

Md Rifat Haidar

Roles: writing-original draft, editing, data collection, implementation

Tasmia Ibrahim

Roles: conceptualization, editing, implementation

Tanjina Rahman

Roles: conceptualization, editing and data management

Mohammad Shahnewaz Sarker

Roles: product development, troubleshooting

Dr. Syada Samsia Noor

Roles: data collection and monitoring

Dr. Shahriar Ahmed

Roles: editing, supervision and troubleshooting

Dr. Mohammad Khaja Mafij Uddin

Roles: supervision and monitoring Dr. Senjuti Kabir

Roles: conceptualization, editing, supervision and troubleshooting

Dr Sayera Banu

Roles: conceptualization, editing, supervision and overall monitoring

## Acknowledgements

We acknowledge the Stop TB partnership for funding and thank to the NTP, local healthcare authorities and the study participants for their contribution.

This research protocol/study was funded by core donors who provide unrestricted support to icddr,b for its operations and research. Current donors providing unrestricted support include the Governments of Bangladesh and Canada. We gratefully acknowledge our core donors for their support and commitment to icddr,b’s research efforts.

## Authors’ information

‘Not applicable’

