## Supporting docs_Manuscript of iDOTS protocol for "A study protocol on assessing the acceptance and effectiveness of a digital adherence technology for TB preventive treatment in Bangladesh": S1 Participant enrollment questionnaire for intervention and control site.docx

**Questionnaire (English)**

**Improving tuberculosis (TB) management at Narsingdi district in Bangladesh: Incorporating TB screening at Integrated Management of Childhood Illness (IMCI) corners and strengthening TB preventive treatment services**

**(Enrolment questionnaire for TPT digital (iDOTS) adherence-Intervention & Control sites)**

| **Hospital name:** | **Interview Date**: ......./......./............ (DD/MM/YY) |
| --- | --- |
| **Interviewer name:** |  |

**Section A: Index Patient History**

| Q. No. | Questions and filters | Categories |
| --- | --- | --- |
| 1. | Participant's index patient Name: |  |
| 2. | TR no: |  |
| 3. | Relationship: |  |
| 4. | Age (in completed years) |  |
| 5. | Gender: |  |

**Section B: Participant’s Demographic Data**

| **Q. No.** | **Questions and filters** | **Categories** |
| --- | --- | --- |
| 6. | Participant 's Name: |  |
| 7. | Gender: | Male  Female  Others |
| 8. | Age (in completed years) |  |
| 9. | Religion | Islam  Hinduism  Christianity  Buddhism  Other |
| 10. | Height/Length | __________(cm) |
| 11. | Weight | __________(kg) |
| 12. | Marital Status  (Select from the list below) | Single  Married  Separated  Divorced  Widow  Other |
| 13. | Major occupation  (Multiple responses) | Farmer, Agriculture  Day labourer  Petty business (hawker)  Small business (small shop owner)  Business  Government Service  Private Service  Rickshaw puller/Van driver  Garments / factory worker  Unemployed / Retired  Professional  Transport worker (Bus, truck, water vehicles etc)  Housewife  Student  Other |
| 14. | Present Address | Road no. . . . . . . . . .  House no: . . . . . . . . .  Vill/Mohallah: . . . . . . . . . . . .  Union: . . . . . . . . . . . .. . . . . . . . .  Sub-District/Thana: . . . . . . . . ..  District: . . . . . . . . . . . . . . . . . .. . |
| 15. | Permanent Address | Road no. . . . . . . . . .  House no: . . . . . . . . .  Vill/Mohallah: . . . . . . . . . . . ...  Union: . . . . . . . . . . . .. . . . . . . . .  Sub-District/Thana: . . . . . . . . ..  District: . . . . . . . . . . . . . . . . . .. . |
| 16. | Mobile no |  |
| 17. | If you have had any formal education, what is the highest class you have passed? | . . . . . . Class |
| 18. | Number of family members (enter in a number) | . . . . . number |
| 19. | Average monthly family income | …………….TK |
| 20. | Average monthly family expenditure | …………….TK |
| 21. | Number of rooms in the household (excluding kitchen) | . . . . . number |
| 22. | What kind of Toilet facility do your household members use? | Flushed Toilet with septic tank  Toilet without septic tank  Improved Pit latrine  open Pit latrine  Hanging toilet  No toilet facility  Other |
| 23. | Do you share the Toilet facility with any other households? | Yes  No  Not known |
| 24. | What is the main source of drinking water for members of your household? | Piped water into dwelling  Piped water to Yard  Public Tap  Tube well  Dug well  Springwater  Rainwater  Surface water (pond/river/lake/canal)  Other |
| 25. | Do you do anything to the water to make it safer to drink? | Yes  No |
| 26. | Does your household have electricity? | Yes  No |
| 27. | Does your household have Solar Electricity | Yes  No |
| 28. | Does your household have the following things? | A television?  A Mobile phone?  A Refrigerator?  An Almira/wardrobe?  A Fan?  A computer/laptop?  Other valuable goods? |
| 29. | What type of fuel does your household mainly use for Cooking? | Electricity  LPG  Natural Gas  Biogas  Kerosene  Charcoal  Wood  Straw/Shrubs  Animal dung  Other  No cooking |
| 30. | Does any member of this household own following things | A car/microbus  An auto bike/CNG  A motorcycle  A bicycle  A rickshaw/Van |
| 31. | Does any member of this household have a bank account? | Yes  No |
| 32. | The main material of the floor? | Brick/Cement/Tiles  Wood/Bamboo  Clay/Earth |
| 33. | The main material of the roof? | Concrete (Brick/Cement/Rod)  Tin /Wood  Clay/Tally/Wood  Straw/Bamboo/Plastic  Others |
| 34. | The main material of the exterior wall? | Brick/Cement  Tin/Wood  Clay/Brick/Wood  Straw/Bamboo/Plastic  Others |

**Section C: Access to Enablers**

| **Q. No.** | **Questions and filters** | **Categories** |
| --- | --- | --- |
| 35. | How many mobile phones do you have in your family | ………. number |
| 36. | Do you have access to or own a mobile phone? | No  Yes, I own a phone that no one else uses  Yes, family shares a phone - I am the primary owner  Yes, family shares a phone - I am NOT the primary owner  Yes, project staff gave me a phone  Other |
| 37. | Active mobile phone type | Basic/Feature phone  Smart phone |
| 38. | Basic operations using the mobile phone  (Multiple responses) | Dial  Receive call  Read message  Write message  Browse internet  Use social networking application  Other |
| 39. | How many hours do you spend on internet browsing in a day? | ………hours |
| 40. | What do you do while browsing internet?  (Multiple responses) | Using social media  Watching videos  Reading Newspapers  Listening to music  Watching television/news  Others |

**Section D: Risk factors / Medical History**

| **Q. No.** | **Questions and filters** | **Categories** |
| --- | --- | --- |
| 41. | BCG vaccination? | Yes  No  Not known |
| 42. | Smoking? | Yes  No  Refused to answer |
| 43. | Other known medical conditions  (Multiple Responses) | Diabetes Mellitus  Hypertension  Asthma  COPD  Other |
| 44. | If “Other”, please specify | . . . . . . . . . . . . . . . . . . . . . . . . . . . . |
