## Supporting docs_Manuscript of iDOTS protocol for "A study protocol on assessing the acceptance and effectiveness of a digital adherence technology for TB preventive treatment in Bangladesh": S2 Pre-intervention survey questionnaire for providers.docx

**Questionnaire (English)**

**Improving tuberculosis (TB) management at Narsingdi district in Bangladesh: Incorporating TB screening at Integrated Management of Childhood Illness (IMCI) corners and strengthening TB preventive treatment services**

**(Pre-intervention questionnaire for TB preventive treatment digital (iDOTS) adherence- Service Provider)**

**Disclosure:** You are invited to this study because you are providing treatment to tuberculosis patients. For those who will have to go through the process of taking TPT medicine, we have proposed a digital monitoring system to make the process easier and more consistent. Your experience and acceptance of the digital monitoring system is very important for our research. There is no major risk/harm involved with you during participating in this study and you will not get any personal benefit. The information you provide to us will help to learn about the acceptance and experiences related to the use of the digital monitoring systems. Your privacy, anonymity will be strictly maintained. You are the only person who can decide about your participation; your participation is entirely voluntary. In addition, you will have the right to withdraw your participation in research at any time and without mentioning any reason.

**Overview of iDOTS:** TPT iDOTS is a software developed by the icddr,b IT team. The primary goal of TPT iDOTS is to enable real-time monitoring of TPT enrolled contacts. The DOTS staff can use the iDOTS dashboard to monitor individual contacts who are taking their daily medication. Every contact will register to iDOTS and he will get a colored box which contain medicine and over the box a phone number is print. Every contacts of a family will get different colored box which are printed with different phone number. Contacts are required to make a call to a specific number to confirm that they have taken their medication. Here in a family can be use same phone number but their calling number are different. Which is determine who are actually calling after taking medicine. After making the call, the contact receives a confirmation SMS from the server, indicating that their call has been recorded successfully.

| **Hospital ID: ☐** | **Participant ID:** ☐☐☐☐ |
| --- | --- |
| **Interviewer ID:** ☐ | **Interview Date**: ......./......./............ (DD/MM/YY) |
| **Study Group:** ☐ |  |

**Section A:** **Participant’s Demographic Data**

| **Q. No.** | **Questions and filters** | **Categories** |
| --- | --- | --- |
| 1. | Participant Name: |  |
| 2. | Sex: | Male  Female  Others |
| 3. | Age (in completed years) |  |
| 4. | Designation |  |
| 5. | Present Address | Road no. . . . . . . . . .  House no :. . . . . . . . .  Vill/Mohallah: . . . . . . . . . . . . . .  Union: . . . . . . . . . . . .. . . . . . . . .  Sub-District/Thana: . . . . . . . . . .  District: . . . . . . . . . . . . . . . . . . . . |
| 6. | Permanent Address | Road no. . . . . . . . . .  House no :. . . . . . . . .  Vill/Mohallah: . . . . . . . . . . . . . .  Union: . . . . . . . . . . . .. . . . . . . . .  Sub-District/Thana: . . . . . . . . . .  District: . . . . . . . . . . . . . . . . . . . . |
| 7. | Years of education (in completed years) | . . . . . . years |
| 8. | Years of experience | . . . . . . years |

**Section B: User experience**

| **Q. No.** | **Performance expectancy** | **Categories** |
| --- | --- | --- |
| 9. | I would find the system useful in my job | Yes  No  Not known |
| 10. | Using the system would improve my job performance | Yes  No  Not known |
| 11. | Using the system would enhance my effectiveness on the job | Yes  No  Not known |
| 12. | Using the system would make it easier  to do my job | Yes  No  Not known |
| 13. | Using the system would improves the quality of the work I do | Yes  No  Not known |
| 14. | Using the system would help better communicate with my patient | Yes  No  Not known |
| 15. | Use of the system can decrease the time needed for my other important job responsibilities | Yes  No  Not known |
|  | **Effort expectancy** |  |
| 16. | It would be easy for me to become skilful at using the system. | Yes  No  Not known |
| 17. | I would find the system easy to use. | Yes  No  Not known |
| 18. | Learning to operate the system would be easy for me. | Yes  No  Not known |
| 19. | I have positive attitude toward using technology | Yes  No  Not known |
| 20. | The system is good | Yes  No  Not known |
| 21. | The system would make work easier | Yes  No  Not known |
| 22. | Working with the system would be better | Yes  No  Not known |
| 23. | The system would help to monitor TPT | Yes  No  Not known |
| 24. | Using the system would help to prepare TPT report | Yes  No  Not known |
| 25. | Using the system would help to understand whether participant taking medicine properly or not | Yes  No  Not known |
| 26. | Using the system would help to remind participants for taking medicine if they don’t take it | Yes  No  Not known |
| 27. | It would take too long to learn how to use the system to make it worth the effort | Yes  No  Not known |
|  | **Social influence** |  |
| 28. | There is necessary arrangement in our hospital to use the system | Yes  No  Not known |
| 29. | In general, the organization would support the use of the system. | Yes  No  Not known |
|  | **Facilitating conditions** |  |
| 30. | I have the resources (phone, balance, network) necessary to use the system. | Yes  No  Not known |
| 31. | I have the technological information necessary to use the system. | Yes  No  Not known |
| 32. | The system is not compatible with other systems I use. | Yes  No  Not known |
| 33. | A specific person (or group) is available for assistance with system difficulties. | Yes  No  Not known |
| 34. | I would have control over using the system | Yes  No  Not known |
| 35. | Given the technology and information it takes to use the system, it would be easy for me to use the system | Yes  No  Not known |
| 36. | Specialized instruction concerning the system would be available to me | Yes  No  Not known |
| 37. | Using the system fits into my work style | Yes  No  Not known |
|  | **Self-efficacy. I could complete a job or task using the system** |  |
| 38. | If my institute ask someone for assistance would be good | Yes  No  Not known |
| 39. | If I can call someone for help if I got stuck | Yes  No  Not known |
| 40. | If I had a lot of time to complete the job for which the system was asked for | Yes  No  Not known |
| 41. | If there was no one around to tell me what to do as I go | Yes  No  Not known |
|  | **Anxiety: The system is somewhat intimidating to me.** |  |
| 42. | I feel apprehensive about using the system. | Yes  No  Not known |
| 43. | I can make mistake while using the system | Yes  No  Not known |
| 44. | I hesitate to use the system for fear of making mistakes I cannot correct. | Yes  No  Not known |
| 45. | Using the system would be somewhat difficult to me | Yes  No  Not known |
|  | **Behavioural intention to use the system** |  |
| 46. | I won´t use the system for my learning | Yes  No  Not known |
| 47. | I would encourage the use of the system for learning to my colleagues | Yes  No  Not known |

*Qualitative guidelines will be prepared based on the results of the quantitative study.
