## Supporting docs_Manuscript of iDOTS protocol for "A study protocol on assessing the acceptance and effectiveness of a digital adherence technology for TB preventive treatment in Bangladesh": S3 Pre-intervention survey qustionnaire for study participants.docx

| **Hospital ID: ☐** | **Participant’s iDOTS ID:** ☐☐☐☐ |
| --- | --- |
| **Interviewer ID:** ☐ | **Interview Date**: ......./......./............ (DD/MM/YY) |

**Section A: Index Patient History**

| Q. No. | Questions and filters | Categories |
| --- | --- | --- |
| 1. | Participant's index patient Name: |  |
| 2. | Relationship: |  |
| 3. | Age (in completed years) |  |
| 4. | Gender: |  |

**Section B: Participant’s Demographic Data**

| **Q. No.** | **Questions and filters** | **Categories** |
| --- | --- | --- |
| 5. | Participant 's Name: |  |
| 6. | Gender: | Male  Female  Others |
| 7. | Age (in completed years) |  |
| 8. | Religion | Islam  Hinduism  Christianity  Buddhism  Other |
| 9. | Height/Length | __________(cm) |
| 10. | Weight | __________(kg) |
| 11. | Marital Status  (Select from the list below) | Single  Married  Separated  Divorced  Widow  Other |
| 12. | Major occupation  (Multiple responses) | Farmer, Agriculture  Day labourer  Petty business (hawker)  Small business (small shop owner)  Business  Government Service  Private Service  Rickshaw puller/Van driver  Garments / factory worker  Unemployed / Retired  Professional  Transport worker (Bus, truck, water vehicles etc)  Housewife  Student  Other |
| 13. | Present Address | Road no. . . . . . . . . .  House no: . . . . . . . . .  Vill/Mohallah: . . . . . . . . . . . .  Union: . . . . . . . . . . . .. . . . . . . . .  Sub-District/Thana: . . . . . . . . ..  District: . . . . . . . . . . . . . . . . . .. . |
| 14. | Permanent Address | Road no. . . . . . . . . .  House no: . . . . . . . . .  Vill/Mohallah: . . . . . . . . . . . ...  Union: . . . . . . . . . . . .. . . . . . . . .  Sub-District/Thana: . . . . . . . . ..  District: . . . . . . . . . . . . . . . . . .. . |
| 15. | Years of education (in completed years) | . . . . . . years [If illiterate code “00”] |
| 16. | Number of family members (enter in a number) | . . . . . number |
| 17. | Average monthly family income | …………….TK |
| 18. | Average monthly family expenditure | …………….TK |
| 19. | Number of rooms in the household (excluding kitchen) | . . . . . number |
| 20. | What kind of Toilet facility do your household members use? | Flushed Toilet with septic tank 1  Toilet without septic tank 2  Improved Pit latrine 3  open Pit latrine 4  Hanging toilet 5  No toilet facility 6  Other 77 |
| 21. | Do you share the Toilet facility with any other households? | Yes  No  Not known |
| 22. | What is the main source of drinking water for members of your household? | Piped water into dwelling 1  Piped water to Yard 2  Public Tap 3  Tube well 4  Dug well 5  Springwater 6  Rainwater 7  Surface water (pond/river/lake/canal) 8  Other 77 |
| 23. | Do you do anything to the water to make it safer to drink? | Yes  No |
| 24. | Does your household have electricity? | Yes  No |
| 25. | Does your household have Solar Electricity | Yes  No |
| 26. | Does your household have the following things? | A television?......................................... 1  A Mobile phone?.................................. 2  A Refrigerator?..................................... 3  An Almira/wardrobe?........................... 4  A Fan?................................................... 5  A computer/laptop?............................... 6  Other valuable goods?........................... 7 |
| 27. | What type of fuel does your household mainly use for Cooking? | Electricity 1  LPG 2  Natural Gas 3  Biogas 4  Kerosene 5  Charcoal 6  Wood 7  Straw/Shrubs 8  Animal dung 9  Other 77  No cooking 0 |
| 28. | Cooking Place? | In the house 1  In a separate house 2  Outdoors… 3 |
| 29. | Does any member of this household own following things | A car/microbus……………………………. 1  An auto bike/CNG………………………… 2  A motor cycle……………………………... 3  A bicycle…………………………………... 4  A rickshaw/Van…………………………… 5 |
| 30. | Does this household own any poultry | Yes  No |
| 31. | Does any member of this household have a bank account? | Yes  No |
| 32. | The main material of the floor? |  |
| 33. | The main material of the roof? |  |
| 34. | The main material of the exterior wall? |  |

**Section C: Access to Enablers**

| **Q. No.** | **Questions and filters** | **Categories** |
| --- | --- | --- |
| 35. | How many mobile phones do you have in your family | ………. number |
| 36. | Do you have access to or own a mobile phone? | No  Yes, I own a phone that no one else uses  Yes, family shares a phone - I am the primary owner  Yes, family shares a phone - I am NOT the primary owner  Yes, project staff gave me a phone  Other |
| 37. | Active mobile phone type | Basic/Feature phone  Smart phone |
| 38. | Basic operations using the mobile phone  (Multiple responses) | Dial  Receive call  Read message  Write message  Browse internet  Use social networking application |
| 39. | How many hours do you spend on internet browsing in a day? | ………hours |
| 40. | What do you do while browsing internet?  (Multiple responses) | Using social media  Watching videos  Reading Newspapers  Listening to music  Watching television/news  Others |

**Section D: Risk factors / Medical History**

| **Q. No.** | | **Questions and filters** | **Categories** |
| --- | --- | --- | --- |
| 41. | BCG vaccination? | | Yes  No  Not known |
| 42. | Smoking? | | Yes  No  Refused to answer |
| 43. | Other known medical conditions  (Multiple Responses) | | Diabetes Mellitus  Hypertension  Asthma  COPD  Other |
| 44. | If “Other”, please specify | | . . . . . . . . . . . . . . . . . . . . . . . . . . . . |

**Section E: User experience**

| **Q. No.** | **Performance expectancy** | **Categories** |
| --- | --- | --- |
| 45. | I would find the system useful | Yes  No  Not known |
| 46. | Using the system would improve my daily medication habit | Yes  No  Not known |
| 47. | Using the system would make it easier  to continue my medication | Yes  No  Not known |
| 48. | Using the system will help me remember when to take my daily medication | Yes  No  Not known |
| 49. | If I finish the medication dose by Using the method, it will prevent me from getting future infections | Yes  No  Not known |
| 50. | Using the system would help better communicate with my providers (Doctors, TLCAs) | Yes  No  Not known |
|  | **Effort expectancy** |  |
| 51. | Learning to using the system is easy for me. | Yes  No  Not known |
| 52. | I would find the system easy to use | Yes  No  Not known |
| 53. | Using the system is good | Yes  No  Not known |
|  | **Facilitating conditions** |  |
| 54. | I have required phone to use the system | Yes  No  Not known |
| 55. | I have enough balance in my phone to use the system | Yes  No  Not known |
| 56. | I have good network coverage to use the system | Yes  No  Not known |
|  | **Anxiety: The system is somewhat difficult to me.** |  |
| 57. | I can make mistake by using the system | Yes  No  Not known |
| 58. | Using the system would be somewhat difficult to me | Yes  No  Not known |
|  | **Behavioural intention to use the system** |  |
| 59. | I won´t use the system | Yes  No  Not known |
| 60. | I would encourage the people living around me for using the system | Yes  No  Not known |

*Qualitative questions will be prepared based on the results of the quantitative study.
