## Supporting docs_Manuscript of iDOTS protocol for "A study protocol on assessing the acceptance and effectiveness of a digital adherence technology for TB preventive treatment in Bangladesh": S4 Post-intervention survey questionnaire for providers.docx

**Section B: User experience**

| **Q. No.** | **Performance expectancy** | **Categories** |
| --- | --- | --- |
| 9. | I found the system useful in my job | Yes  No  Not known |
| 10. | Using the system enables me to accomplish tasks more quickly | Yes  No  Not known |
| 11. | Using the system increases my productivity | Yes  No  Not known |
| 12. | Using the system improved my job performance | Yes  No  Not known |
| 13. | Using the system enhanced my effectiveness on the job | Yes  No  Not known |
| 14. | Using the system makes it easier  to do my job | Yes  No  Not known |
| 15. | Using the system improved the quality of the work I do | Yes  No  Not known |
| 16. | Using the system helps better communicate with my patient | Yes  No  Not known |
| 17. | Use of the system had no effect on the performance of my job | Yes  No  Not known |
| 18. | Use of the system decreased the time needed for my important job responsibilities | Yes  No  Not known |
|  | **Effort expectancy** |  |
| 19. | It was easy for me to become skilful at using the system. | Yes  No  Not known |
| 20. | I found the system easy to use. | Yes  No  Not known |
| 21. | Learning to operate the system was easy for me. | Yes  No  Not known |
| 22. | I had positive attitude toward using technology | Yes  No  Not known |
| 23. | Using the system was a good idea | Yes  No  Not known |
| 24. | The system made work easier | Yes  No  Not known |
| 25. | Working with the system was better | Yes  No  Not known |
| 26. | The system was helpful to monitor TPT | Yes  No  Not known |
| 27. | Using the system helped to prepare TPT report | Yes  No  Not known |
| 28. | Using the system helped to understand whether participant taking medicine properly or not | Yes  No  Not known |
| 29. | Using the system helped to remind participants for taking medicine if they don’t take it | Yes  No  Not known |
| 30. | It takes too long to learn how to use the system to make it worth the effort | Yes  No  Not known |
|  | **Social influence** |  |
| 31. | My higher authority instructed me to use the system | Yes  No  Not known |
| 32. | People who are working here using the system thought that I should use the system | Yes  No  Not known |
| 33. | There was necessary arrangement in our hospital to use the system | Yes  No  Not known |
| 34. | In general, the organization supported the use of the system. | Yes  No  Not known |
|  | **Facilitating conditions** |  |
| 35. | I had the resources (phone, balance, network) necessary to use the system. | Yes  No  Not known |
| 36. | I have had the information necessary to use the system. | Yes  No  Not known |
| 37. | The system was not compatible with other systems I use. | Yes  No  Not known |
| 38. | A specific person (or group) was available for assistance with system difficulties. | Yes  No  Not known |
| 39. | I had control over using the system | Yes  No  Not known |
| 40. | Given the technology and information it takes to use the system, it would be easy for me to use the system | Yes  No  Not known |
| 41. | Specialized instruction concerning the system was available to me | Yes  No  Not known |
| 42. | Using the system fits into my work style | Yes  No  Not known |
|  | **Self-efficacy. I could complete a job or task using the system** |  |
| 43. | If my institute ask someone for assistance could be good | Yes  No  Not known |
| 44. | If I could call someone for help if I got stuck | Yes  No  Not known |
| 45. | If I had a lot of time to complete the job for which the software was provided | Yes  No  Not known |
| 46. | If there was no one around to tell me what to do as I go | Yes  No  Not known |
|  | **Anxiety: The system is somewhat intimidating to me.** |  |
| 47. | I felt apprehensive about using the system. | Yes  No  Not known |
| 48. | It scared me to think that I could lose a lot of information using the system by hitting the wrong key. | Yes  No  Not known |
| 49. | I hesitated to use the system for fear of making mistakes I cannot correct. | Yes  No  Not known |
| 50. | The system was somewhat intimidating to me | Yes  No  Not known |
|  | **Behavioural intention to use the system** |  |
| 51. | I won´t use the system for my learning in future | Yes  No  Not known |
| 52. | I recommended the use of the system for learning to my colleagues | Yes  No  Not known |

*Qualitative guidelines will be prepared based on the results of the quantitative study.
