## Supporting docs_Manuscript of iDOTS protocol for "A study protocol on assessing the acceptance and effectiveness of a digital adherence technology for TB preventive treatment in Bangladesh": S5 Post-intervention survey questionnaire for study participants.docx

| **Hospital ID:** | **Participant iDOTS ID:…………………………..** |
| --- | --- |
| **Interviewer ID:** | **Interview Date**: ......./......./............ (DD/MM/YY) |
| **Mobile no:** |  |

**Section A: User experience**

| **Q. No.** | **Performance expectancy** | **Categories** |
| --- | --- | --- |
| 1. | I found the system useful | Yes  No  Not known |
| 2. | Using the system enabled me to finish my day to day medication | Yes  No  Not known |
| 3. | Using the system increases my intention to take care of myself | Yes  No  Not known |
| 4. | Using the system improved my daily medication habit | Yes  No  Not known |
| 5. | Using the system made it easier  to continue my medication | Yes  No  Not known |
| 6. | Using the system helped me to remember the time of taking medication | Yes  No  Not known |
| 7. | Using the system helped to finish my medication and increased my chances of not getting the disease | Yes  No  Not known |
| 8. | Using the system helped better communicate with my providers (Doctors, TLCAs) | Yes  No  Not known |
|  | **Effort expectancy** |  |
| 9. | Learning to using the system was easy for me. | Yes  No  Not known |
| 10. | I found the system easy to use | Yes  No  Not known |
| 11. | Using the system is good | Yes  No  Not known |
| 12. | I liked using the system. | Yes  No  Not known |
| 13. | I found the system to be flexible to interact with | Yes  No  Not known |
| 14. | Using the system takes too much time from my normal duties | Yes  No  Not known |
|  | **Facilitating conditions** |  |
| 15. | I had required phone to use the system | Yes  No  Not known |
| 16. | I had enough balance in my phone to use the system | Yes  No  Not known |
| 17. | I had good network coverage to use the system | Yes  No  Not known |
|  | **Anxiety: The system is somewhat intimidating to me.** |  |
| 18. | I made mistake by using the system | Yes  No  Not known |
| 19. | Using the system was somewhat difficult to me | Yes  No  Not known |
|  | **Behavioural intention to use the system** |  |
| 20. | I didn’t want to use the system | Yes  No  Not known |
| 21. | I recommended the use of system for learning to people living around me | Yes  No  Not known |

*Qualitative guidelines will be prepared based on the results of the quantitative study.
